# Empirical evidence for safety of mechanical ventilation during simulated cardiopulmonary resuscitation on a physical model

**DOI:** 10.1101/2020.04.27.20081778

**Authors:** Leonardo Bugarin de Andrade Neumamm, Alcendino Cândido Jardim-Neto, Gabriel Casulari da Motta-Ribeiro

**Affiliations:** Pulmonary Engineering Laboratory, Biomedical Engineering Program, Alberto Luiz Coimbra Institute for Graduate Studies and Research in Engineering (COPPE), Federal University of Rio de Janeiro, Rio de Janeiro, Brazil; Marcílio Dias Navy Hospital, Rio de Janeiro, Brazil; AmericanCor Hospital, Rio de Janeiro, Brazil; Lung Mechanics Laboratory, Department of Biomedical Engineering, Stevens Institute of Technology, Hoboken, USA

**Keywords:** Cardiopulmonary resuscitation, Ventilatory modes, Mechanical ventilation, Chest compressions

## Abstract

**Background:** Cardiac arrest is a critical event requiring adequate and timely response in order to increase patient’s chance of survival. In patients mechanically ventilated with advance airways cardiopulmonary resuscitation (CPR) maneuver may be simplified by keeping the ventilator on. This work assessed the response of a intensive care mechanical ventilator to CPR using a patient manikin ventilated in three conventional modes.

**Methods:** Volume controlled (VCV), pressure controlled (PCV) and pressure regulated volume controlled (PRVC) ventilation were applied in a thorax physical model with or without chest compressions. The mechanical ventilator was set with inspiratory time of 1.0 s, ventilation rate of 10 breaths/minute, positive end-expiratory pressure of 0 cmH2O, FiO2 of 1.0, target tidal volume of 600 ml and trigger level of -20 cmH2O. Airway opening pressure and ventilatory flow signals were continuously recorded..

**Results:** Chest compression resulted in increased airway peak pressure in all ventilation modes (p<0.001), specially with VCV (137% in VCV, 83% in PCV, 80% in PRVC). However, these pressures were limited to levels similar to release valves in manual resuscitators (∼60 cmH_2_O). In pressure controlled modes tidal/minute volumes decreased (PRVC=11%, p=0.027 and PCV=12%, p<0.001), while still within the variability observed during bag-valve-mask ventilation. During VCV, variation in tidal/minute volumes were not significant (p=0.140). Respiratory rates were constant with and without chest compression.

**Conclusions:** A intensive care mechanical ventilator could provide adequate ventilation during compressions in a manikin model while using conventional ventilation modes.

## 1. INTRODUCTION

Cardiac arrest is a critical event in a intensive care unit, requiring fast and coordinated response in order to increase patient’s chance of survival^1^. To revert that condition, cardiopulmonary resuscitation (CPR) should be performed to achieve return of spontaneous circulation. Although the American Heart Association (AHA) determines that circulation must be prioritized during CPR, ventilation should be considered^2^. This guidelines recommend a CPR maneuver with 5 to 6 cm chest compressions at a rate of 100 to 120 per minute without interruptions for ventilation, which should be provided at 10 breathings per minute if an advanced airway is placed. Additionally, it has been suggested that tidal volumes of 6 to 7 ml/kg are adequate during CPR although visually perceived chest movements during inflation are considered sufficient to ensure satisfactory ventilation^3^.

In approximately one third of CPR maneuvers performed in intensive care units, patients are already under mechanical ventilation^4^ and disconnection to change for a bag-valve device may delay the begin of CPR-ventilation. There are ventilatory modes able to synchronize ventilation to chest compressions^5,6^, yet these are not available on most intensive care unit ventilators; which still rely on standard ventilatory modes. Therefore, the aim of this work was to evaluate the ventilation provided by a commercial ventilator during simulated CPR maneuver on a human physical model.

## 2. METHODS

### 2.1. STUDY DESIGN

A CPR maneuver was simulated in a patient manikin suitable for chest compression (SimMan^®^, Laerdal Medical, Norway) intubated and mechanically ventilated (Servo-i, Dräger, Germany). Chest compressions were performed at a rate of 100 per minute by one of the authors, an experienced health professional who was blind to the ventilatory mode in execution. Effective compressions were achieved with the aid of a metronome for frequency accuracy and feedback from the model’s sensor for compressions’ depth.

Mechanical ventilation was set to mimic recommended parameters for bag-valve-mask ventilation during CPR: positive end-expiratory pressure = 0 cmH_2_O, respiratory rate = 10 breaths per minute, fraction of inspired oxygen = 1.0, inspiratory time per cycle = 1.0 s. Trigger pressure was set to the least sensible value (-20 cmH_2_O) and peak pressure alarm was set to the maximum possible value (120 cmH_2_O). Three ventilatory modes were tested: volume-controlled (VCV), pressure-controlled (PCV) and pressure-regulated volume-controlled ventilation (PRVC). During VCV and PRVC modes, tidal volume was set to 600 mL; while on PCV mode the targeted inspiratory pressure was adjusted to achieve 600 mL tidal volume prior to compressions, resulting in 16 cmH_2_O.

Ventilatory modes were tested in random order and triplicate sessions. Each session was comprised of 5 minutes ventilation without chest compressions followed, uninterrupted, by 2 minutes of ventilation with chest compressions.

### 2.2. DATA COLLECTION

During each CPR maneuver airway pressure and ventilatory flow were continuously measured at the outer end of the endotracheal tube (ID 8.0) with a purpose-built data acquisition system. The system was composed of two differential pressure transducers (143PC01D and 176PC07HD2, Honeywell, USA), a variable orifice pneumotachograph (PN279331 Hamilton-Medical, Switzerland) and a dedicated signal filter and amplifier. All sensors were calibrated before the start of the experiment and signals were recorded with a software written in LabView (National Instruments, USA). Tidal volumes were obtained by numerical integration of the flow signals within each semi-automatically detected breath cycle.

### 2.3. STATISTICAL ANALYSIS

Data are presented as median, first and third quartiles. Tidal volumes, inspiratory peak flow, peak inspiratory pressure, inspiratory time and minute ventilation were compared among ventilatory modes (PCV *vs*. VCV *vs*. PRVC) by using Kruskal-Wallis test and Dunn-Sidak correction for multiple comparisons, and within ventilatory modes (compression vs. no compression), with Wilcoxon rank sum test. Signal and statistical analysis were performed in Matlab (MathWorks, USA) and significance level was set at 0.05.

## 3. RESULTS

The tested ventilation modes responded differently to CPR maneuver. Manual chest compressions were performed at a rate of 101.9 [99.0 109.9] per minute (median [first and third quartiles], for all modes combined) without modifying the respiratory rate with the chosen trigger level (Figure 1A). VCV had larger variations in pressure compared to PCV and PRVC, while pressure controlled modes showed larger variations in inspiratory volume, compared to VCV (Table 1).

**Table 1.**
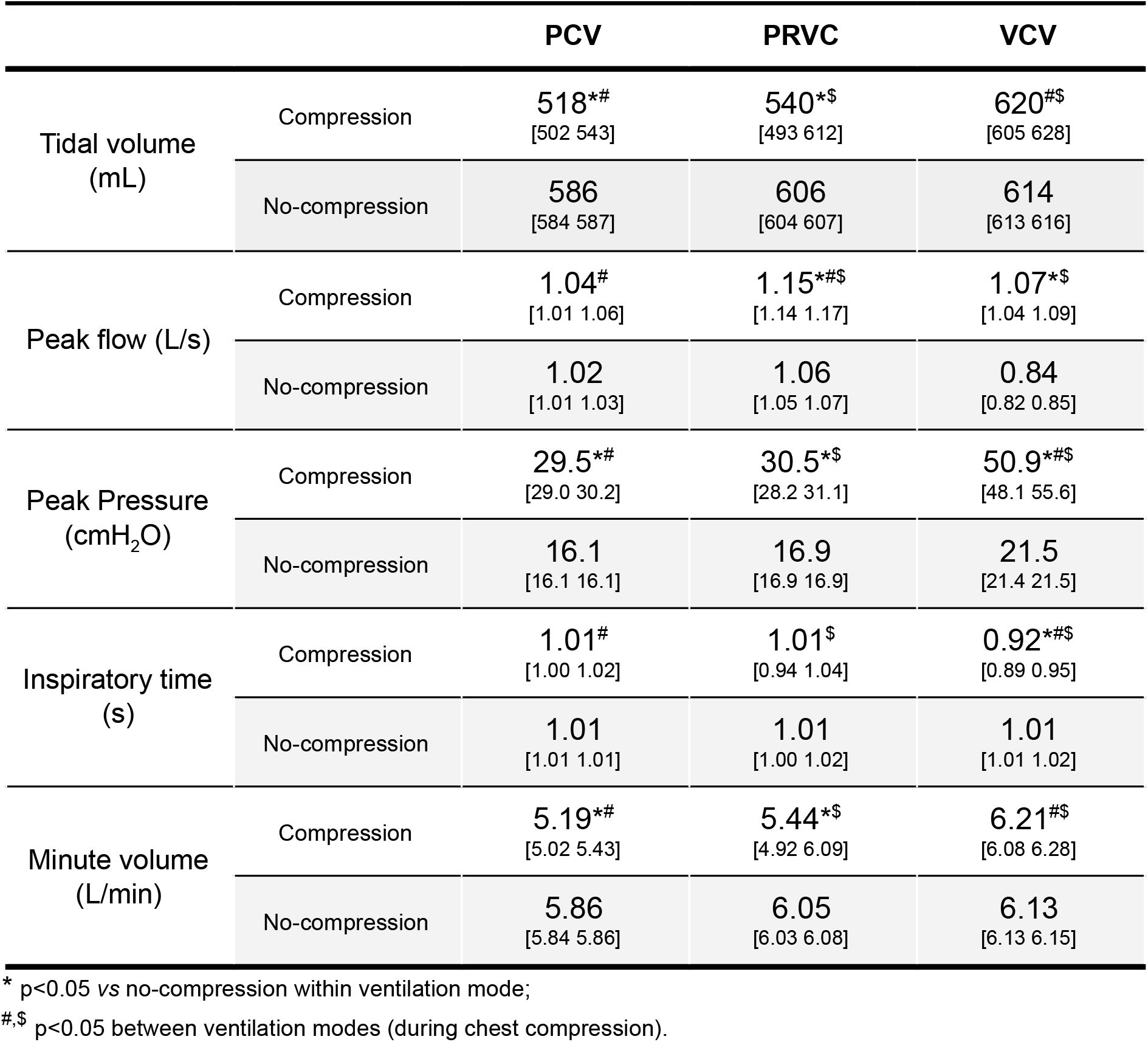
Ventilatory data from ventilation mode with and without chest compression (median [first and third quartiles]).

**Figure 1.**
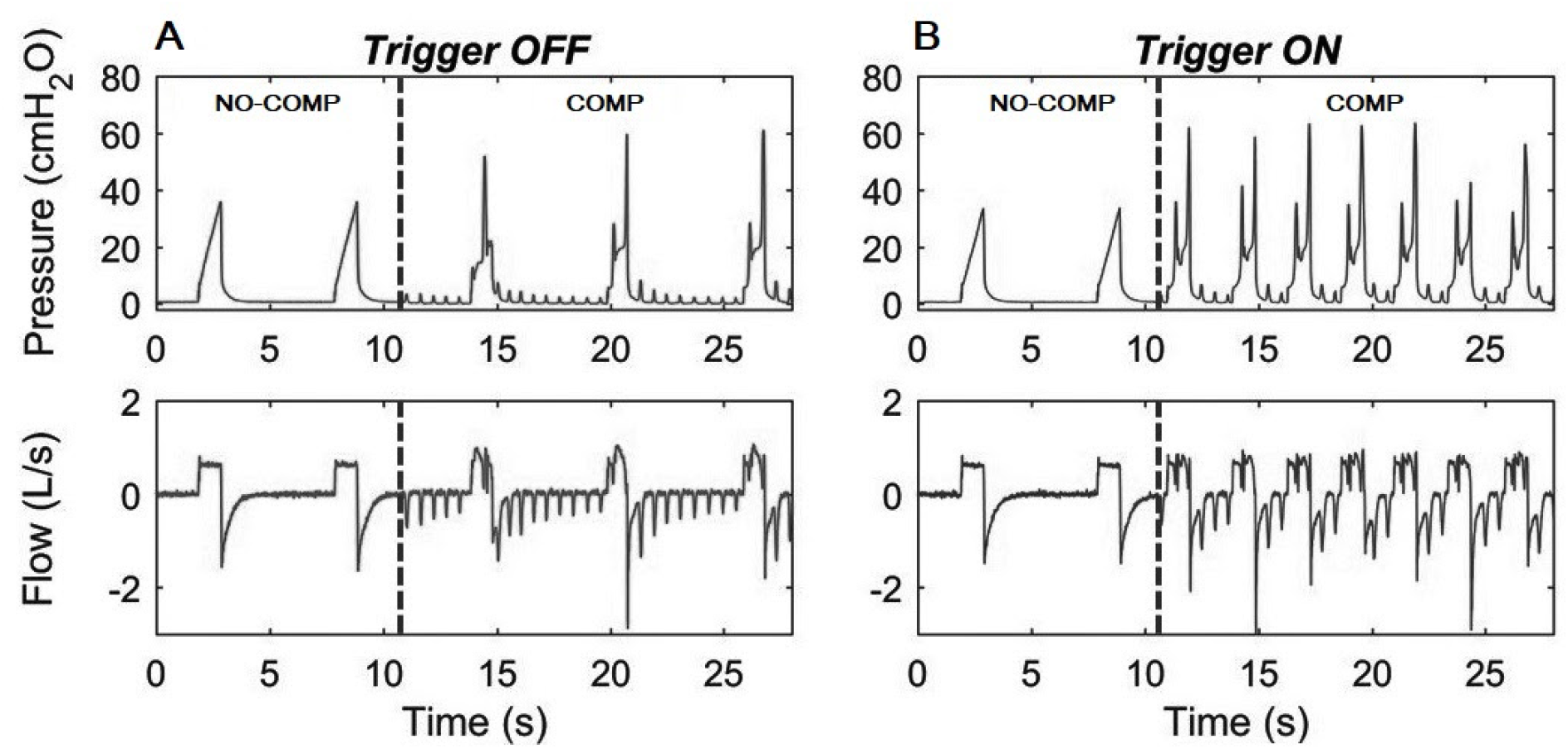
No-compression (NO-COMP) to compression (COMP) transition (vertical dashed lines) during volume-controlled ventilation with the ventilator trigger pressure set to a high sensibility (“Trigger ON” column) and low sensibility (“Trigger OFF” column) conditions. Without compression the pressure and flow had typical waveforms. The compressions with “active trigger” (ON) were sufficient to initiate premature cycles, increasing the respiratory rate in comparison to the “inactive trigger” (OFF) in which the respiratory rate remained similar to pre-compression condition.

Chest compressions increased peak inspiratory pressure in all ventilation modes (83% in PCV, 80% in PRVC and 137% in VCV, p<0.001; Table 1 and Figure 2). Tidal and minute volume were maintained in VCV (Figure 2) with a decrease in inspiratory time compared to no-compression (Table 1). In contrast, tidal volume decreased in PRVC (11%, p=0.027) and PCV (12%, p<0.001), resulting in decreased minute volume, without changes in inspiratory time. Peak flow, during CPR, increased only in PRVC and VCV (p<0.001).

**Figure 2.**
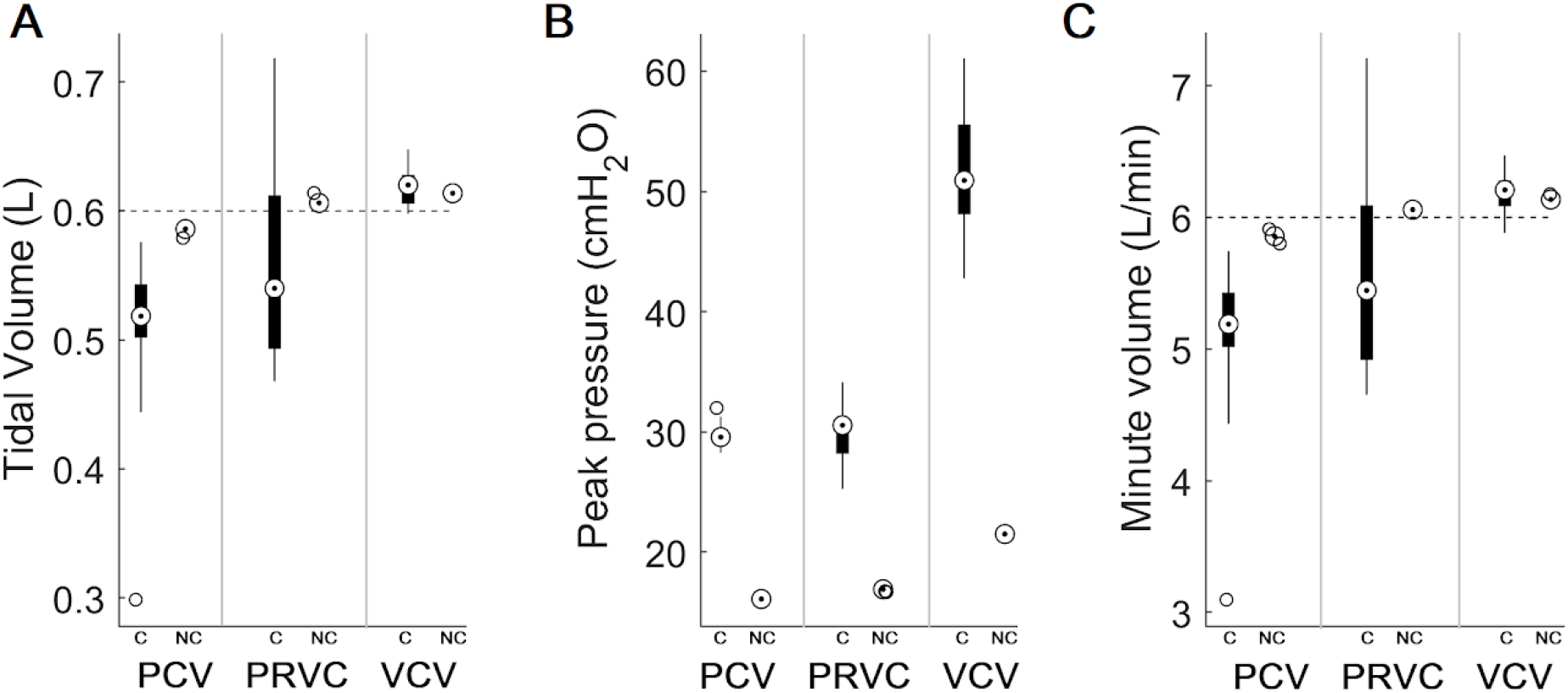
Ventilatory data from all cycles obtained with (C) and without (NC) chest compressions for each ventilation mode: PCV (Pressure-controlled ventilation, blue), PRVC (Pressure-regulated volume controlled, red) and VCV (Volume-controlled ventilation, green). Dashed black lines indicates recommended values by the consensus. Stars show statistically significant difference between C and NC conditions.

## 4. DISCUSSION

Although it is established that there is no need to disconnect the patient to provide manual ventilation during CPR if an advanced airway is already placed and connected to a mechanical ventilator^7,8^, there has been little information on the effects of different ventilatory modes under such circumstance. Our work main findings were the increased airway peak pressure in all ventilation modes, specially with VCV, and the reduction in tidal volumes with PRVC and PCV during simulated CPR maneuver.

We hypothesize that the interaction between chest compression forces, ventilator valves and control algorithms resulted in the measured increase in airway pressure and different responses among ventilation modes. The sudden application of external opposing forces to lung inflation would emulate an increase in respiratory system elastance, increasing airway pressures for fixed inspiratory volumes or decreasing those volumes for constant inspiratory pressure, as observed. Nevertheless, the highest peak pressures during VCV were about 60 cmH2O within the pressure set for safety release valves in manual resuscitators, and the variability in tidal volumes were within the observed during mask-bag-valve ventilation^9^. Thus, our data suggests that, in a CPR scenario, none of the ventilation modes would result in clinically prohibitive pressures and would deliver volumes acceptable by the current standards.

The regularity of the inspiratory times obtained during CPR maneuver indicates the overall absence of premature ventilatory cycles triggered by chest compression forces. The least sensible trigger pressure on the ventilator settings (-20 cmH_2_O) was effective in avoiding those undesired cycles, maintaining the respiratory rate similar to pre-compression condition^10^. This is of considerable importance since it has been shown that hyperventilation during CPR^11^ may lead to higher intrathoracic pressures, decreased coronary perfusion pressures, induce hypotension and consequently death^12,13^

This work presented limitations regarding the physical model, ventilation strategie and CPR maneuver. We used a stable unicompartimental lung model without the temporal and compression-related mechanical changes expected on a real human lung under the represented conditions. Also, a single ventilator and parameters set were tested during compressions performed by a single individual. In order to mitigate those limitations, we assessed both the ventilator performance prior to the simulated CPR maneuver, using a calibrated data acquisition system independent from the ventilator, and the compression rate using an accelerometer. By doing that, we might have minimized chances of measuring effects other than changes in ventilation profile due ventilator-chest-compression interactions. *In vivo* studies are need to validate effectiveness of ventilation.

As our data indicates that the maintenance of mechanical ventilation during CPR maneuvers is not clinically prohibitive, this could lead to a more efficient use of human resources in the intensive care environment by eliminating the need of a dedicated health professional to maintain ventilation during CPR through alternative methods as the bag-valve, in already mechanically ventilated patients. This is of particular importance during crisis scenarios where there are few health professionals and when aerosolization of infectious diseases may be a concern.

## 5. CONCLUSION

Conventional ventilation modes during simulated CPR maneuvers did not resulted in prohibitive airway peak pressures and tidal volumes, while providing adequate minute ventilation. Thus, disconnection from mechanical ventilator may be disregarded even when compression synchronized modes are not available.

## Data Availability

All experimental data is available under request.

## 7. ACKNOWLEDGEMENTS

The authors thankfully acknowledge the Instituto de Pesquisas Biomédicas of the Marcílio Dias Navy Hospital (Rio de Janeiro, Brazil) for providing physical space and material resources for the experiment, and Prof. D.Sc Antonio Giannella-Neto for his valuable intellectual support to this work. This work was partially supported by the Brazilian National Council for Scientific and Technological Development (CNPq), Coordination of Superior Level Staff Improvement (CAPES) and Carlos Chagas Filho Foundation for Research Support of the State of Rio de Janeiro (FAPERJ) with study grants to the authors and funding to acquire the mechanical ventilator used. Sponsors had no role on study design, data collection and analysis, text writing, or decision about publication.

